# Stepped care for young people at clinical high risk for psychosis: a real-world study

**DOI:** 10.64898/2026.02.05.26345683

**Authors:** Alexander Broekhuijse, Abhishek Saxena, Barbara Walsh, Catalina Mourgues-Codern, Hesham Mukhtar, Samantha Howard, Scott W. Woods, Albert R. Powers, Emily A. Farina

## Abstract

**Objective:** Despite recommendations that young people at clinical high risk (CHR) for psychosis receive stepped treatment, few programs have published details of their clinical models or outcomes. This study describes the preliminary effectiveness of a risk calculator-informed stepped care model used at the Yale PRIME Clinic, a specialized outpatient clinic for young people at CHR.

**Methods:** Seventy-one individuals (ages 12-25) at CHR enrolled in Yale’s PRIME Clinic during the first four years of the treatment program. Participants completed clinical assessments at six timepoints over two years of treatment within a care model informed by an empirically grounded psychosis risk calculator. Linear mixed-effect models were fit to examine changes in clinical symptoms over time, and sensitivity analyses evaluated differences in clinical trajectories between completers and non-completers.

**Results:** Individuals engaged in treatment demonstrated significant and sustained improvements in positive, negative, general, disorganized, and depressive symptoms. Improvements in positive symptoms emerged by 6 months and continued to improve across most subsequent timepoints (6, 12, and 24 months). Pattern mixture analyses suggested that clinical trajectories did not significantly differ between completers and non-completers, though non-completers possessed more heterogeneous trajectories.

**Conclusions:** A stepped care model informed by individualized risk calculator scores was feasible for delivery in a specialized outpatient setting, and was associated with broad symptom improvement for young people at CHR. Further controlled studies with blinded raters are needed to further confirm the efficacy of stepped care models and isolate the active components of treatment.

**Highlights:**

- Participants at clinical high risk for psychosis experienced significant reductions in attenuated psychotic symptoms and improvements in mood while enrolled in a risk-calculator-informed stepped care treatment model.
- Participants who disengaged from treatment did not have significantly different clinical trajectories than those who remained in care.
- The results suggest preliminary evidence for the feasibility of implementing a risk-calculator-informed stepped care model.

## Introduction

Psychotic disorders are disabling conditions associated with adverse individual, family, and socioeconomic impacts (1,2). Well-validated instruments to detect clinical high risk for psychosis (CHR) syndromes enable earlier identification of individuals at risk for psychosis (3–5), potentially shortening the duration of untreated psychosis or preventing onset altogether (6). Young people at CHR are at elevated risk for developing a psychotic disorder (7) in addition to experiencing distressing symptoms and functional declines warranting treatment (8,9). Several interventions show promise (10–14) though no single intervention has yet demonstrated consistent or durable effects (15).

Given the absence of treatment options, experts recommend stepped care (SC), a tiered system of treatments of varying intensity which calibrates treatment intensity to a person’s current level of need (16). This approach may be especially beneficial for the heterogeneous presentations of CHR (17–19), in part because reserving higher-intensity interventions, such as antipsychotic medications, for those with the greatest need avoids unnecessary treatment exposure, while the provision of adjunctive care can target common comorbidities.

Support for SC was formally recognized in the United States beginning in 2018, when the Substance Abuse and Mental Health Services Administration (SAMHSA) funded programs nationwide to design, implement, and evaluate SC models for CHR (20–23). Preliminary work demonstrated strong rationale and promising outcomes for SC (16,22). However, the efficacy of SC remains unconfirmed, highlighting the need for additional research examining novel SC models.

The Yale PRIME Clinic developed a SC model guided by the North American Prodrome Longitudinal Study (NAPLS) Risk Calculator (24), a validated prognostic tool that quantifies an individual’s likelihood of developing a psychotic disorder within two years. In this model, the risk calculator informs stepping decisions to tailor treatment intensity to individual risk level.

Here, we evaluate the preliminary effectiveness of this SC model across two years of treatment in a specialized setting. Consistent with standard handling of missing data in clinical trial research (25,26), we examine factors associated with treatment discontinuation alongside differences in clinical trajectory and variability between those who completed a full two years of treatment and those who did not. We hypothesized that: (1) participants in treatment would display a significant decrease in attenuated psychotic symptoms and depressive symptoms over time; (2) participants who disengaged from care would display similar average symptom trajectories to those who remained in care; and (3) despite similar means, participants who discontinued early would have significantly more variable symptom trajectories than those who remained in care, mirroring findings in large scale naturalistic psychotherapy research (27,28).

## Methods

### Participants

Between 2018 and 2022, 71 treatment-seeking individuals across Connecticut enrolled in the PRIME Clinic within the Yale School of Medicine in New Haven, Connecticut. Inclusion criteria were (1) ages 12-25 years, (2) IQ≥70, (3) fluency in English, (4) ability to provide informed consent, and (5) met criteria for a CHR syndrome based on the Structured Interview for Psychosis Risk Syndromes (SIPS) (4) with active symptoms. Participants were largely referred from the community through the clinic’s established referral network (local schools, healthcare providers, and social services), and less frequently self-referrals. This study was approved by the Yale University Institutional Review Board (IRB Approval#: 2000024266).

### The PRIME Stepped Care Model

The PRIME SC model is comprised of three steps (**Figure 1**), providing evidence-informed psychosocial interventions in initial steps and reserving antipsychotic medication (AP) for those with the highest symptom burden and conversion risk. ’Adjunctive care’ was available across all steps, including pharmacotherapy for comorbidities, peer, group, and family support, substance use risk reduction, and vocational and/or educational support.

**Figure 1.**
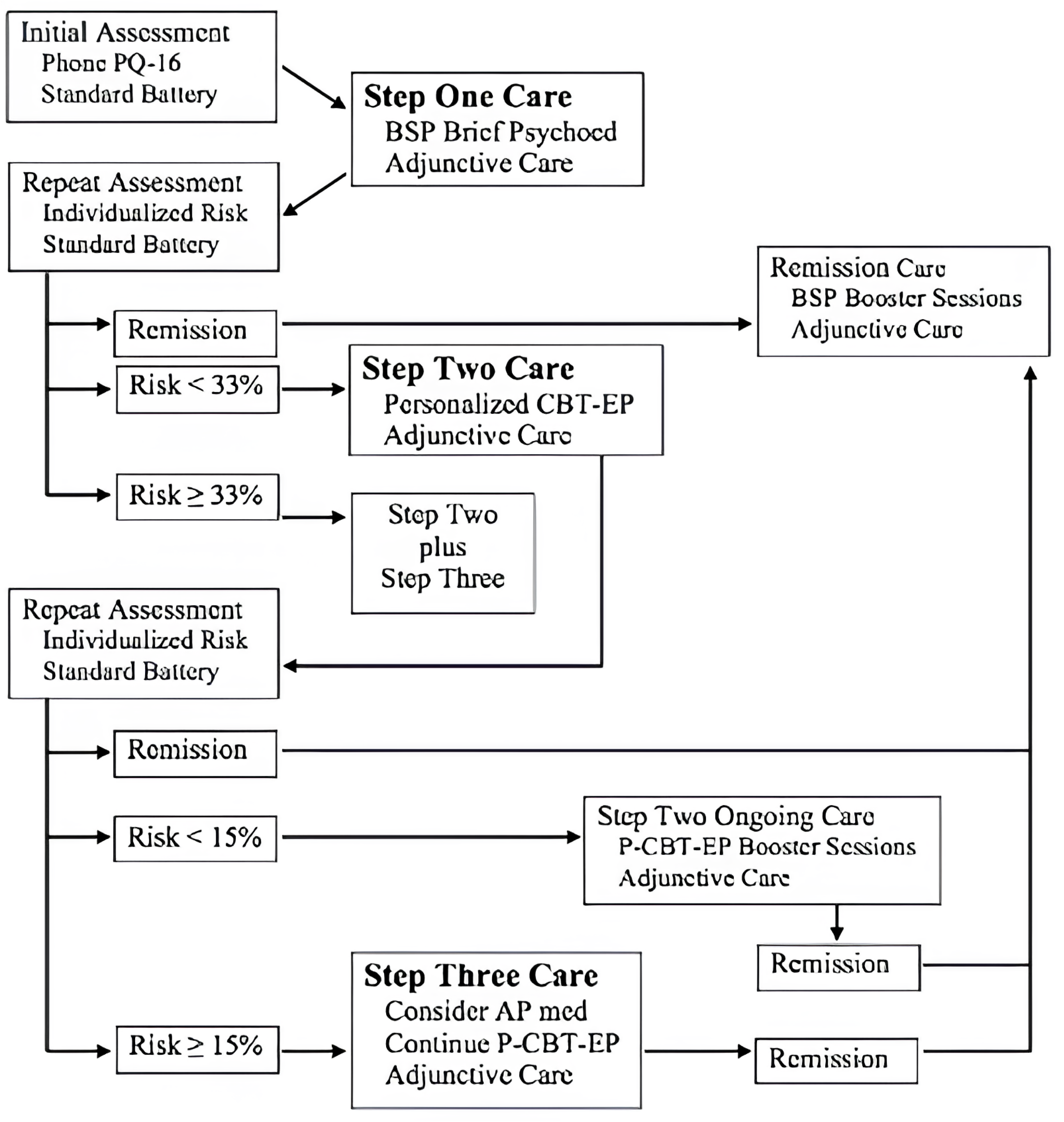
**The stepped care model for CHR employed at the PRIME clinic.** *Alt Text:* Flowchart showing a three-step stepped care model with risk-based decision points determining treatment escalation or remission pathways.

### Implementation details are in the Online Supplement

Individualized risk estimates using the NAPLS risk calculator (24) were produced at baseline, following Step One, and every six months during care, and were used alongside client preferences and clinical judgment to inform stepping decisions. All participants began with Step One. Following Step One, participants with a risk score <33% proceeded to Step Two; those with a score >33% proceeded directly to Step Three. At all subsequent six-month assessments, a 15% threshold governed step assignment: participants scoring >15% were assigned to Step Three, while those scoring <15% received Step Two. Participants could step down as well as up if risk scores changed. These cutoffs were drawn from existing research on two-year psychosis transition rates (7,29): 33% identifies the upper tertile of risk warranting immediate intensive intervention and aligns with meta-analytic two-year conversion estimates (∼25%), while 15% marks the point within the risk score distribution at which converters begin to outnumber non-converters in the original sample (29).

#### Step One: Brief Supportive Psychoeducation (BSP)

All participants began treatment with BSP, a flexible psychoeducational intervention for CHR originally utilized in the NAPLS studies (30). Content included education about CHR symptoms, the vulnerability-stress model, the role of the clinician and family, problem-solving, coping strategies, and identification of personalized risk and protective factors. BSP was delivered over three sessions within two months of baseline.

#### Step Two: Personalized CBT for Early Psychosis (P-CBT-EP)

Individuals who reached Step Two were offered P-CBT-EP, a flexible manualized cognitive-behavioral intervention tailored to early psychosis symptoms (manuals available upon reasonable request). P-CBT-EP was intended to be delivered in 15 sessions over approximately six months. Content included the relation between thinking, feeling, and behavior, cognitive distortions, automatic negative thoughts, probability overestimation, jumping to conclusions, and exposure. Content was selected based on a shared formulation of problematic areas developed collaboratively with participants. While P-CBT-EP attends to positive symptoms, its content is derived from core cognitive-behavioral theory and could be adapted for participants meeting CHR criteria for familial risk and functional decline alone. Participants continued to work with their assigned psychotherapist from Step One across all steps. While other psychotherapies have been examined to address APS since the start of this study in 2018, none have been confirmed as efficacious for improving APS and thus the development and testing of new interventions was warranted (15,31).

All participants who completed Step One received Step Two. Those with a risk score <33% received Step Two alone; those with a risk score ≥ 33% after Step One BSP and/or significantly distressing CHR symptoms were offered both Step Two and Step Three.

After completion of Step Two care, participants achieving CHR full remission (six consecutive months with all positive symptoms scored at a two or less on the SIPS (32)) following P-CBT-EP were offered remission care, consisting of in-person booster sessions (minimum every six months), telephone monitoring (minimum every two months), and adjunctive care as needed. Participants who were not in remission but whose two-year risk of psychosis is < 15% proceeded to P-CBT-EP booster sessions and adjunctive care. Lastly, participants who were not in remission, but whose two-year risk of psychosis was ≥ than 15% proceeded to Step Three.

#### Step Three: Consideration of Antipsychotic Medication

Participants who reached Step Three were offered antipsychotic medication. Choice of antipsychotic medication was based on client preference related to adverse event profile, and dosing was based on the minimum dose that sustains remission with minimal emergent adverse effects. More information regarding medical decision making can be found in the Online Supplement.

In addition to Step Three care, each participant met with a psychiatrist at baseline and every 6 months, with increased frequency based on clinical need. Participants who enrolled already taking an antipsychotic entered treatment on Step 3 but still received BSP followed by P-CBT-EP.

### Assessments

Clinical status and functioning were assessed at baseline, and 2, 6, 12, 18, and 24 months. Participants also received as-needed assessment if conversion to psychosis was suspected. Assessments administered included a sociodemographic questionnaire, all instruments included in the risk calculator (24), the Columbia Suicide Severity Rating Scale (33), the Structured Interview for Psychosis Risk Syndromes, Patient Health Questionnaire, 9 (PHQ-9) (34), and SAMHSA’s National Outcome Measures (NOMs (35)). Assessments were completed by each participant’s psychotherapist. Clinicians were certified in the administration and interpretation of the SIPS through a two-day certification process through Yale University, and all individual components of the NAPLS risk calculator. Details on all assessments and administration protocols can be found in the Online Supplement.

The SIPS (4) is a structured clinical interview composed of four scales that measure positive symptoms (range 0-30), negative symptoms (range 0-36), disorganized communication (range 0-24), and general psychopathology (range 0-24). Each item is scored on a scale of 0 (Absent) to 6 (Psychotic Level Severity), with a 6 on the positive items indicating the presence of frank psychosis and a 3-5 indicating CHR-level severity. A comprehensive review of studies utilizing the SIPS suggests an average total positive symptom scores of 9.9, while nationwide longitudinal research suggests averages of 11.92 (negative), 5.15 (disorganized), and 9.19 (general) for individuals at CHR (36,37). CHR status was identified using the full criteria of prodromal symptoms as detailed in the SIPS. The SIPS was administered at screening pre-baseline to ascertain CHR status and then again at predetermined intervals as a component for the risk calculator assessment.

The Patient Health Questionnaire-9 (PHQ-9; (34)) is a 9-item self-report measure assessing the frequency of DSM depressive symptoms over the past two weeks, rated on a 4-point scale from 0 (*not at all*) to 3 (*nearly every day*). Total scores range from 0 to 27, with established cutoffs of 5, 10, 15, and 20 corresponding to mild, moderate, moderately severe, and severe depression.

The NAPLS psychosis risk calculator (24) is an individualized prognostic tool derived from the NAPLS-2 cohort which utilizes six assessments to estimate one- and two-year transition to psychosis risk for individuals that meet CHR criteria. Details on specific assessments and administration protocol can be found in the Online Supplement.

### Statistical analysis

Statistical analyses were conducted in R version 4.5.1. Descriptive analyses examined participant sociodemographic and clinical characteristics. Mean symptom severities were compared to those found in prior large CHR samples (36,38,39) and established cutoffs (40). Linear mixed effects models were fit in accordance with the intent-to-treat principle to examine outcomes (SIPS and PHQ-9 scores), adjusted for age and sex, with time as a categorical fixed effect and a random intercept for each client (REML; Satterthwaite degrees of freedom). Intraclass-correlation and fixed/conditional R^2^ values were calculated to evaluate model fit. Pairwise contrasts (Tukey) compared individual timepoints. Linear mixed-effects models enabled the use of all available measurements regardless of missing observations under a Missing-At-Random (MAR) assumption (41). All data for included covariates were complete.

To examine whether clinical trajectories differed by retention and disengagement, pattern mixture analyses were run as Missing-Not-At-Random (MNAR) sensitivity analyses.

Participants were classified by last observed visit (completers, disengagement at 6, 12, 18, and 24 months). Within each cutoff, linear mixed-effects models were fit with timepoint, pattern (reached, did not reach), and their interaction as fixed effects, with participant as a random effect. To evaluate whether early treatment discontinuation prior to two years was associated with greater heterogeneity in clinical outcomes, homogeneity of variance across completers and non-completers at 12, 18, and 24 months was compared using the Brown-Forsythe test (median centred Levene).

## Results

### Participants

**Table 1** includes baseline sociodemographic and clinical characteristics. Of the 71 individuals enrolled, 58% were male. Average age was 15.6 years old (SD=2.69), and participants were 53.5% White, 35.2% Black, 4.2% Asian, 2.8% Indigenous American/Alaskan, and 4.2% “Other” unlisted race. Eight participants were prescribed antipsychotic medication prior to enrolment, while 20 were prescribed antidepressant medication. During treatment, 20 additional participants were prescribed antipsychotic and/or antidepressant medication.

**Table 1.** Demographic and Baseline Clinical Characteristics.

| Baseline Characteristic | Overall | Percentage | Male mean | Female mean |
| --- | --- | --- | --- | --- |
| Age (years) | 15.61 ± 2.69 | | 15.29 | 16.07 |
| *Sex* |  |  |  |  |
| Male | 41 | 57.7% |  |  |
| Female | 30 | 42.3% |  |  |
| *Race* |  |  |  |  |
| White | 38 | 53.5% |  |  |
| Black | 25 | 35.2% |  |  |
| Asian | 3 | 4.2% |  |  |
| Indigenous American | 2 | 2.8% |  |  |
| Other | 3 | 4.2% |  |  |
| *Clinical Characteristics* |  |  |  |  |
| Positive symptoms (baseline) | 13.89 ± 3.03 | | 14.29 | 13.33 |
| General symptoms (baseline) | 12.87 ± 3.35 | | 13.46 | 12.03 |
| Negative symptoms (baseline) | 12.99 ± 4.50 | | 12.98 | 13.00 |
| Disorganized symptoms (baseline) | 5.36 ± 2.30 | | 5.88 | 4.62 |
| Patient Health Questionnaire, 9 (baseline) | 13.81 ± 5.99 | | 14.56 | 12.81 |

*Overall column shows mean ± SD; Male/Female columns show means.*

At baseline, participants presented with above-average positive (M=13.9, SD=3.03), negative (M=13.0, SD=4.50), and general (M=12.9, SD=3.35) symptoms, average disorganized (M=5.36, SD=2.30) symptoms, as well as moderate depressive symptom severity (PHQ-9; M=13.81, SD=5.99). Average estimates of conversion risk were 20.03% (SD=11.63) for one-year and a 25.66% (SD=14.35) for two-years.

The full two-year protocol was completed by 25 participants (35.2%), 12 (16.9%) were discharged due to full symptom remission (32), 10 (14.1%) were discharged due to repeated no-shows, 5 (7%) were discharged due to conversion to psychosis, and 19 (26.8%) voluntarily withdrew from treatment. Participants who left treatment, including those discharged due to symptom remission, were not reevaluated. Of those who completed two years of treatment, 45% achieved full remission by the 24-month assessment. Attrition occurred gradually across timepoints, with 71 participants at baseline, 54 (76%) participants at two months, 43 (60%) participants at six months, 40 (56%) participants at 12 months, 36 (51%) participants at 18 months, and 26 (37%) participants at 24 months. Overall, 71 participants participated in Step 1, 54 participants participated in Step 2, and 20 participants participated in Step 3. While allowed, no participants moved backwards between steps in practice.

### H1: Change in participant clinical outcomes

To evaluate symptom change, linear mixed-effects models were run on 267 observations across 71 participants. An intraclass correlation coefficient was calculated to identify the total variance attributable to between-participant differences. Intraclass correlations suggested moderate clustering within participants and moderate within-person similarity among repeated measures (Positive symptoms=0.46, Negative symptoms=0.54, General symptoms=0.44, Disorganization symptoms=0.42, PHQ-9=0.48).

Symptom trajectories are visualized in **Figure 2**. Timepoint effects were significant for all symptom domains, including positive (F=78.40, df=5 and 204, p<0.001), negative (F=26.40, df=5 and 171, p<0.001), disorganized (F=7.55, df=5 and 174, p<0.001), general (F=29.89, df=5, 177, p<0.001), and depressive symptom severity (F=27.02, df=5 and 196, p<0.001).

**Figure 2.**
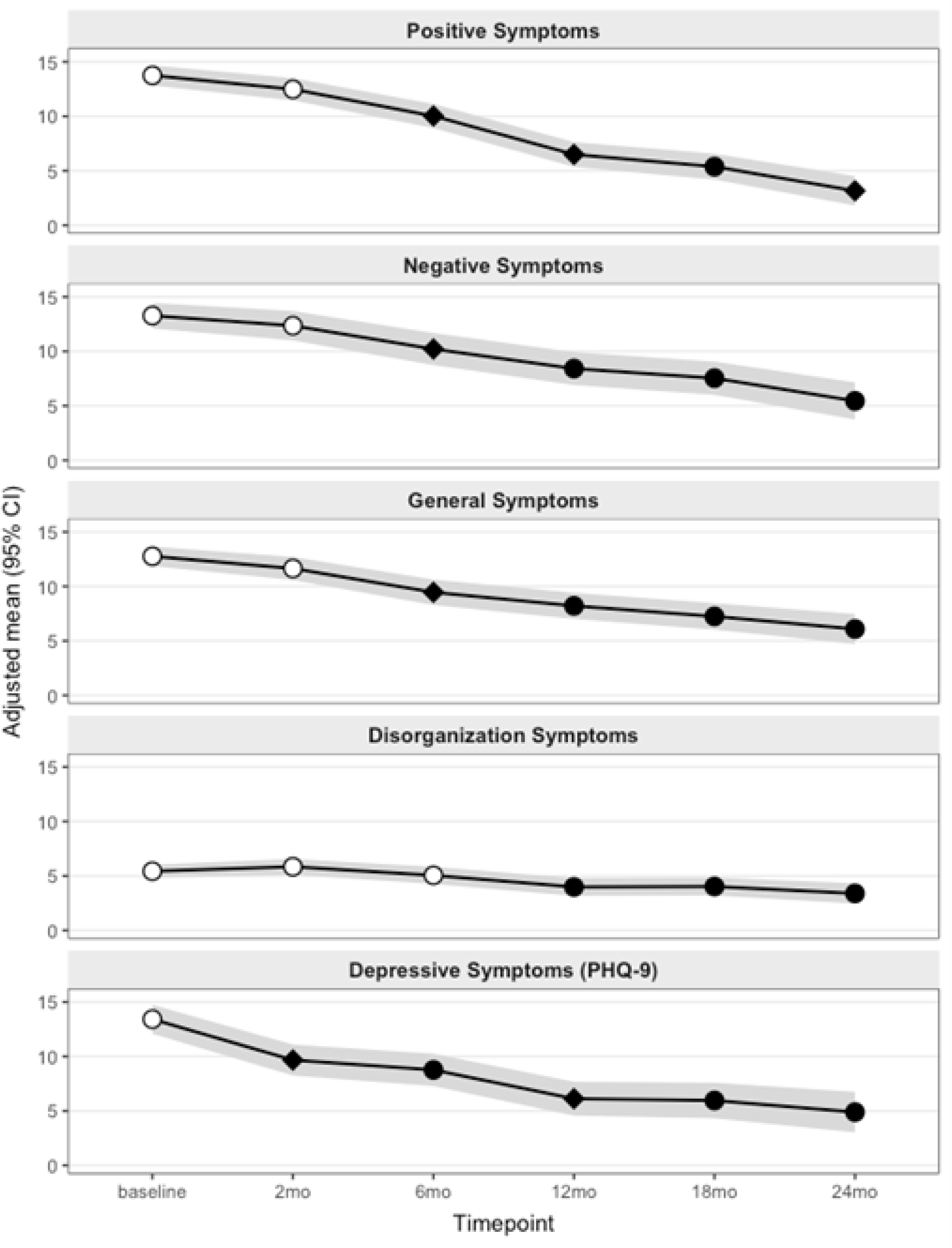
Adjusted SIPS scores by timepoint (baseline to 24 months). Linear mixed effects models adjusted for age and sex. Points are estimated marginal means, and shading indicates 95% confidence intervals. Positive (P), negative, (N), general (G), and disorganized (D) symptoms are each represented by an individual line. A ○ indicates no significant change, a ● indicates significant change compared to baseline, and a ♦ indicates both a significant change compared to baseline and compared to the previous measurement point. *Alt text:* Five-panel line graph showing symptom trajectory EMMs with 95% CIs across five SIPS symptom domains from baseline to 24 months.

Estimated marginal means (EMMs) declined from baseline to 24 months for all outcomes, with significant baseline contrasts at multiple timepoints after multiplicity control. At 12 months, participants showed significant reductions in positive (Δ=7.25, p<0.001), negative (Δ=4.86, p<0.001), general (Δ=4.55, p<0.001), disorganized (Δ=1.45, p=.011), and depressive symptoms (Δ=7.29, p<0.001). Positive (Δ=3.34, p<0.001), negative (Δ=2.97, p=0.018), and general (Δ=2.12, p=0.05) symptoms continued improving through 24 months, while disorganized and depressive symptoms stabilized. Changes in positive and general symptoms were significant at 6 months; disorganized symptoms by 12 months; and depressive symptoms by 2 months.

Fixed effects accounted for 48% of variance in positive symptoms (marginal R²=0.48), increasing to 72% when including random effects (conditional R²=0.72). Other symptom domains possessed greater between-person heterogeneity (N=0.25→0.65; G=0.31→0.61; D=0.12→0.49; PHQ-9=0.28→0.62). Model summaries and EMMs with 95% CIs are reported in Table 2 (Holm vs. Baseline); timepoint-wise contrasts (Tukey all-by-all) are in the online supplement.

**Table 2.** Changes in estimated SIPS and PHQ-9 scores from baseline to each subsequent measurement point.

| Outcome | Timepoint | n | EMM ± SE | 95% CI (EMM) | Δ vs baseline† | 95% CI (Δ) | t(df) | p (Holm) | F (timepoint omnibus) | p (omnibus) |
| --- | --- | --- | --- | --- | --- | --- | --- | --- | --- | --- |
| P | baseline | 71 | 13.75 ± 0.47 | [12.82, 14.68] |  |  |  |  | F(5, 204) = 78.40 | <.001 |
|  | 2mo | 54 | 12.48 ± 0.52 | [11.46, 13.51] | -1.27 | [-2.65, 0.11] | -2.39(204) | 0.018 |  |  |
|  | 6mo | 43 | 10.03 ± 0.56 | [8.92, 11.13] | -3.72 | [-5.21, -2.24] | -6.51(206) | <.001 |  |  |
|  | 12mo | 40 | 6.50 ± 0.58 | [5.36, 7.64] | -7.25 | [-8.78, -5.72] | -12.33(208) | <.001 |  |  |
|  | 18mo | 36 | 5.38 ± 0.60 | [4.19, 6.57] | -8.37 | [-9.96, -6.78] | -13.69(209) | <.001 |  |  |
|  | 24mo | 26 | 3.16 ± 0.69 | [1.81, 4.51] | -10.59 | [-12.39, -8.79] | -15.32(209) | <.001 |  |  |
| G | baseline | 70 | 12.76 ± 0.46 | [11.86, 13.66] |  |  |  |  | F(5, 178) = 29.89 | <.001 |
|  | 2mo | 45 | 11.64 ± 0.54 | [10.58, 12.70] | -1.12 | [-2.56, 0.32] | -2.03(178) | 0.044 |  |  |
|  | 6mo | 34 | 9.46 ± 0.59 | [8.29, 10.63] | -3.30 | [-4.89, -1.72] | -5.44(180) | <.001 |  |  |
|  | 12mo | 32 | 8.21 ± 0.61 | [7.01, 9.41] | -4.55 | [-6.17, -2.93] | -7.31(180) | <.001 |  |  |
|  | 18mo | 31 | 7.24 ± 0.62 | [6.03, 8.46] | -5.52 | [-7.16, -3.88] | -8.75(180) | <.001 |  |  |
|  | 24mo | 23 | 6.09 ± 0.70 | [4.71, 7.46] | -6.67 | [-8.52, -4.83] | -9.43(181) | <.001 |  |  |
| N | baseline | 70 | 13.27 ± 0.59 | [12.10, 14.45] |  |  |  |  | F(5, 171) = 26.40 | <.001 |
|  | 2mo | 45 | 12.35 ± 0.68 | [11.00, 13.70] | -0.92 | [-2.62, 0.78] | -1.41(171) | 0.160 |  |  |
|  | 6mo | 34 | 10.21 ± 0.75 | [8.74, 11.69] | -3.06 | [-4.93, -1.19] | -4.26(173) | <.001 |  |  |
|  | 12mo | 32 | 8.42 ± 0.76 | [6.91, 9.93] | -4.86 | [-6.77, -2.94] | -6.59(173) | <.001 |  |  |
|  | 18mo | 31 | 7.53 ± 0.77 | [6.01, 9.06] | -5.74 | [-7.68, -3.80] | -7.70(173) | <.001 |  |  |
|  | 24mo | 23 | 5.45 ± 0.86 | [3.74, 7.15] | -7.83 | [-10.01, -5.65] | -9.35(173) | <.001 |  |  |
| D | baseline | 70 | 5.43 ± 0.31 | [4.82, 6.03] |  |  |  |  | F(5, 174) = 7.55 | <.001 |
|  | 2mo | 45 | 5.84 ± 0.36 | [5.13, 6.56] | 0.41 | [-0.57, 1.40] | 1.09(174) | 0.551 |  |  |
|  | 6mo | 34 | 5.05 ± 0.40 | [4.26, 5.84] | -0.38 | [-1.46, 0.70] | -0.92(177) | 0.551 |  |  |
|  | 12mo | 32 | 3.98 ± 0.41 | [3.17, 4.80] | -1.45 | [-2.55, -0.34] | -3.40(177) | 0.003 |  |  |
|  | 18mo | 31 | 4.03 ± 0.42 | [3.20, 4.85] | -1.40 | [-2.52, -0.28] | -3.26(177) | 0.004 |  |  |
|  | 24mo | 23 | 3.39 ± 0.47 | [2.46, 4.33] | -2.04 | [-3.29, -0.78] | -4.22(178) | <.001 |  |  |
| PHQ-9 | baseline | 63 | 13.42 ± 0.67 | [12.09, 14.75] |  |  |  |  | F(5, 196) = 27.02 | <.001 |
|  | 2mo | 51 | 9.67 ± 0.72 | [8.25, 11.10] | -3.75 | [-5.68, -1.81] | -5.04(197) | <.001 |  |  |
|  | 6mo | 45 | 8.76 ± 0.75 | [7.28, 10.25] | -4.65 | [-6.67, -2.64] | -6.00(199) | <.001 |  |  |
|  | 12mo | 40 | 6.12 ± 0.79 | [4.57, 7.68] | -7.29 | [-9.40, -5.19] | -9.01(202) | <.001 |  |  |
|  | 18mo | 35 | 5.96 ± 0.83 | [4.33, 7.59] | -7.46 | [-9.67, -5.25] | -8.78(202) | <.001 |  |  |
|  | 24mo | 25 | 4.89 ± 0.94 | [3.03, 6.75] | -8.53 | [-11.02, -6.03] | -8.88(202) | <.001 |  |  |

† Δ vs baseline; Holm-adjusted p-values. R<sup>2</sup>(marginal/conditional) and ICC by outcome are reported in text.

### H2: Difference in trajectory by disengagement timepoint

Pattern-mixture sensitivity analyses were used to examine whether clinical change varied by disengagement status. Across symptom domains, interaction effects (timepoint x pattern) were mostly non-significant, providing no evidence that clinical symptoms separated completers from non-completers. Two exceptions were noted for negative symptoms at the 6-month (F=6.46, df=2 and 49, p=0.014) and 12-month (F=5.73, df=2 and 82, p<0.01) assessments, while non-completers showed lower negative symptoms. Otherwise, completers and non-completers demonstrated statistically similar improvements across treatment. Group-level trajectories between completers and non-completers and individual and mean participant change scores are visualized in the online supplement.

### H3: Variability in symptom trajectory by retention status

Brown-Forsythe tests indicated heteroscedasticity for positive symptoms (F=9.48, df=2 and 10, p<0.001) and PHQ-9 scores (F=4.94, df= 2 and 11, p<0.001), and homoscedasticity for negative symptoms, general symptoms, and disorganized symptoms. As such, non-completers had significantly more variability in their positive symptom and PHQ-9 scores than completers.

## Discussion

In an outpatient sample of young people at CHR, a risk-calculator-informed SC model was feasible and associated with sizable improvements in attenuated psychotic and depressive symptoms over two years. Main findings were that 1) participants demonstrated significant and sustained improvements in positive, negative, general, disorganized, and depressive symptoms across two years of SC; 2) symptom trajectories were comparable between treatment completers and non-completers, suggesting early disengagement was not primarily driven by clinical worsening, and 3) symptom variability was greater among non-completers, indicating more heterogeneous patterns of change in those who discontinued care early.

Importantly, 56% of participants remained enrolled in care for at least one year of treatment, and the full completion rate was comparable to rates in general outpatient mental health care in the United States, psychosocial treatment trials for CHR, and existing SC trials (16,42,43).

Findings demonstrate that a SC model informed by empirically derived risk estimates can be successfully delivered in a specialized outpatient environment with meaningful clinical improvement. Consistent reductions across symptom domains, particularly in positive and depressive symptoms, indicate that this model was associated with transdiagnostic symptom relief. Results align with prior research showing significant within-subject improvement among youths at CHR engaged in cognitive-behavioral or psychoeducational interventions (44). By 12 months, 25% of participants had achieved full remission, rising to 36.6% at 18 months and 50% by 24 months, comparable to documented remission rates in observational studies and existing trials of SC (27.2% in a recent randomized controlled trial of staged treatment) (16,45). The observed conversion rate (7.04%) was significantly below calculated risk estimates at baseline (25%; exact binomial test, p < .001). However, the calculator was developed in an observational cohort, and its calibration in treatment seeking samples remains understudied (46); the discrepancy may reflect any combination of treatment effects or underlying population differences. Additionally, clinician feedback indicated it was easily usable, and assessments offered valuable treatment insights. While causative associations cannot be made as this is an uncontrolled pre-post study, these findings are promising and lay the foundation for future work in controlled settings.

Nonsignificant differences in clinical trajectories between completers and non-completers suggest that early treatment disengagement was not likely driven by symptom exacerbation or treatment ineffectiveness. Clinician observations (described in Supplement), suggest improvements in functioning over time may have driven disengagement, as many participants chose to discharge early to engage in social, academic, and occupational activities. Consistent with these observations, fewer negative symptoms were observed in non-completers.

However, disengagement could indicate that care did not meet the needs of an individual, and future qualitative work should seek to understand service user perspectives on specific SC models.

This study adds to the limited evidence base describing the implementation and outcomes of SC for CHR. Prior work indicated promising outcomes for SC for reducing attenuated positive symptoms and functioning (20,22), and the present study demonstrates similar benefits. Notably, the present study further extends the evidence base by testing a novel SC model informed by a psychosis risk calculator that also demonstrated improvements in negative, general, disorganization, and depressive symptoms (20,21,47).

Interpretation should consider several limitations. First, as an uncontrolled pre-post study, improvements could reflect regression to the mean, naturalistic improvement, or concurrent treatment effects. Second, data on intervention fidelity, the relative contributions of individual steps, adjunctive care, and patient satisfaction were not collected in a systematic or standardized way, and this data should be prioritized in future trials evaluating SC. Third, assessments were conducted by participants’ psychotherapists, raising the possibility of allegiance or expectancy effects. Fourth, missing data limited potential analyses, including analyses of patient functioning. Fifth, this model was implemented in a specialty clinic with extensive psychosis risk training and its feasibility in settings with more limited training, or in typical community practice settings, remains unclear. Future randomized controlled trials to confirm efficacy and trials examining implementation factors and effectiveness broadly are needed. Sixth, eight participants entered the study with previous antipsychotic and antidepressant exposure, which could have influenced their response to treatment. Seventh, heterogeneous reasons for disengagement may have influenced sensitivity analyses.

## Conclusion

A SC model for young people at CHR, informed by empirically derived conversion risk estimates, is implementable in specialized settings and associated with broad sustained improvement across symptom domains. Integrating individualized risk prediction with SC bridges the gap between empirically informed precision risk assessment and clinical practice.

Further research in randomized controlled trials is needed to confirm the efficacy of SC and to evaluate the relative contributions of individual steps and specific stepping decisions.

## Data Availability

Data is available upon request to first author.

## Acknowledgements

This work was funded by SAMHSA Grant Number: 6H79SM081190-01M001. Additionally, this work was funded in part by the State of Connecticut, Department of Mental Health and Addiction Services, but this publication does not express the views of the Department of Mental Health and Addiction Services or the State of Connecticut. The views and opinions expressed are those of the authors.

## References

1. Eaton WW: The burden of mental disorders. Epidemiol Rev 30: 1–14, 2008.

2. Arias D, Saxena S, Verguet S: Quantifying the global burden of mental disorders and their economic value. EClinicalMedicine 54: 101675, 2022.

3. Woods SW, Addington J, Cadenhead KS, et al.: Validity of the Prodromal Risk Syndrome for First Psychosis: Findings From the North American Prodrome Longitudinal Study. Schizophrenia Bulletin 35: 894–908, 2009.

4. Miller TJ, McGlashan TH, Rosen JL: Prodromal assessment with the Structured Interview for Prodromal Syndromes and the Scale of Prodromal Symptoms: predictive validity, interrater reliability, and training to reliability. Schizophrenia Bulletin 29: 703–715, 2003.

5. Woods SW, Lympus C, McGlashan TH, et al.: The Mini-SIPS: development of a brief clinical structured interview guide to diagnosing DSM-5 Attenuated Psychosis Syndrome and training outcomes. BMC Psychiatry 22: 784, 2022.

6. Fusar-Poli P, McGorry PD, Kane JM: Improving outcomes of first-episode psychosis: an overview. World Psychiatry 16: 251–265, 2017.

7. Salazar De Pablo G, Radua J, Pereira J, et al.: Probability of Transition to Psychosis in Individuals at Clinical High Risk: An Updated Meta-analysis. JAMA Psychiatry 78: 970, 2021.

8. Haining K, Gajwani R, Gross J, et al.: Clinical and Functional Outcomes of Community-Recruited Individuals at Clinical High-Risk for Psychosis: Results From the Youth Mental Health Risk and Resilience Study (YouR-Study). Schizophrenia Bulletin Open 5: sgae029, 2024.

9. Fusar-Poli P, Rocchetti M, Sardella A, et al.: Disorder, not just state of risk: Meta-analysis of functioning and quality of life in people at high risk of psychosis. Br J Psychiatry 207: 198–206, 2015.

10. Fusar-Poli P: Preventive Treatments for Psychosis: Umbrella Review (Just the Evidence. Front Psychiatry 10: 764, 2019.

11. Zheng Y, Xu T, Zhu Y, et al.: Cognitive Behavioral Therapy for Prodromal Stage of Psychosis—Outcomes for Transition, Functioning, Distress, and Quality of Life: A Systematic Review and Meta-analysis. Schizophrenia Bulletin 48: 8–19, 2022.

12. Herrera SN, Sarac C, Phili A, et al.: Psychoeducation for individuals at clinical high risk for psychosis: A scoping review. Schizophrenia Research 252: 148–158, 2023.

13. Woods SW, Addington J, Bearden CE, et al.: Psychotropic medication use in youth at high risk for psychosis: Comparison of baseline data from two research cohorts 1998–2005 and 2008–2011. Schizophrenia Research 148: 99–104, 2013.

14. Mukhtar H, Zhou D, Farina EA, et al.: Prediction of antipsychotic medication inception in antipsychotic-naive youth at clinical high risk for psychosis. Psychol Med 55: e241, 2025.

15. Minichino A, Davies C, Karpenko O, et al.: Preventing psychosis in people at clinical high risk: an updated meta-analysis by the World Psychiatric Association Preventive Psychiatry section. Mol Psychiatry 30: 2773–2782, 2025.

16. McGorry PD, Mei C, Amminger GP, et al.: A Sequential Adaptive Intervention Strategy Targeting Remission and Functional Recovery in Young People at Ultrahigh Risk of Psychosis: The Staged Treatment in Early Psychosis (STEP) Sequential Multiple Assignment Randomized Trial. JAMA Psychiatry 80: 875, 2023.

17. Solmi M, Soardo L, Kaur S: Meta-analytic prevalence of comorbid mental disorders in individuals at clinical high risk of psychosis: the case for transdiagnostic assessment. Molecular Psychiatry 28: 2291–2300, 2023.

18. Shah JL, Jones N, Van Os J, et al.: Early intervention service systems for youth mental health: integrating pluripotentiality, clinical staging, and transdiagnostic lessons from early psychosis. The Lancet Psychiatry 9: 413–422, 2022.

19. Addington J, Piskulic D, Liu L, et al.: Comorbid diagnoses for youth at clinical high risk of psychosis. Schizophrenia Research 190: 90–95, 2017.

20. Cosgrove JA, Crocker LD, Breitborde NJK, et al.: Stepped Care Interventions for Psychosis Risk: Findings From Clinical High Risk for Psychosis Grantee Programs. PS appi.ps.20240505, 2025.

21. Shapiro DI, Grattan RE, Laquidara JR, et al.: Feasibility of a SteppedLCare Intervention for Those at Clinical High Risk for Psychosis in the United States. Early Intervention Psych 20: e70120, 2026.

22. Hamilton SA: Symptomatic and Functional Outcomes Among Individuals at High Risk for Psychosis Participating in Stepped Care. Psychiatr Serv appips20230188, 2023.

23. Moe AM, Cowan HR, Manges M, et al.: The influence of complex psychiatric comorbidities on treatment for clinical high risk for psychosis: A preliminary study. Early Intervention Psych 18: 888–893, 2024.

24. Cannon TD, Yu C, Addington J: An individualized risk calculator for research in prodromal psychosis. American Journal of Psychiatry 173: 980–988, 2016.

25. Little RJ, D’Agostino R, Cohen ML, et al.: The Prevention and Treatment of Missing Data in Clinical Trials. N Engl J Med 367: 1355–1360, 2012.

26. The Prevention and Treatment of Missing Data in Clinical Trials. Washington, D.C., National Academies Press, 2010 [cited 2026 Feb 24]Available from: https://www.nationalacademies.org/publications/12955

27. Zieve GG, Persons JB, Yu LAD: The Relationship Between Dropout and Outcome in Naturalistic Cognitive Behavior Therapy. Behavior Therapy 50: 189–199, 2019.

28. Lopes RT, Gonçalves MM, Sinai D, et al.: Clinical outcomes of psychotherapy dropouts: does dropping out of psychotherapy necessarily mean failure? Rev Bras Psiquiatr 40: 123–127, 2017.

29. Carrión RE, Cornblatt BA, Burton CZ, et al.: Personalized Prediction of Psychosis: External Validation of the NAPLS-2 Psychosis Risk Calculator With the EDIPPP Project. AJP 173: 989–996, 2016.

30. Miklowitz DJ, O’Brien MP, Schlosser DA, et al.: Family-Focused Treatment for Adolescents and Young Adults at High Risk for Psychosis: Results of a Randomized Trial. Journal of the American Academy of Child & Adolescent Psychiatry 53: 848–858, 2014.

31. Zheng Y, Xu T, Zhu Y, et al.: Cognitive Behavioral Therapy for Prodromal Stage of Psychosis—Outcomes for Transition, Functioning, Distress, and Quality of Life: A Systematic Review and Meta-analysis. Schizophrenia Bulletin 48: 8–19, 2022.

32. Woods SW, Walsh BC, Addington J, et al.: Current status specifiers for patients at clinical high risk for psychosis. Schizophrenia Research 158: 69–75, 2014.

33. Posner K, Brown GK, Stanley B, et al.: The Columbia–Suicide Severity Rating Scale: Initial Validity and Internal Consistency Findings From Three Multisite Studies With Adolescents and Adults. AJP 168: 1266–1277, 2011.

34. Kroenke K, Spitzer RL, Williams JBW: The PHQ-9: Validity of a brief depression severity measure. J Gen Intern Med 16: 606–613, 2001.

35. Riske-Morris M, Hussey DL, Olgac T, et al.: Commentary on the Revision of SAMHSA’s National Outcome Measures (NOMs) for Discretionary Programs. J Behav Health Serv Res 51: 302–308, 2024.

36. Addington J, Liu L, Buchy L, et al.: North American Prodrome Longitudinal Study (NAPLS 2): The Prodromal Symptoms. Journal of Nervous & Mental Disease 203: 328–335, 2015.

37. Woods SW, Walsh BC, Powers III AR, et al.: Reliability, Validity, Epidemiology, and Cultural Variation of the Structured Interview for Psychosis-Risk Syndromes (SIPS) and the Scale of Psychosis-Risk Symptoms (SOPS); in Handbook of Attenuated Psychosis Syndrome Across Cultures. Edited by Li H, Shapiro DI, Seidman LJ, et al. Springer Nature Switzerland AG, 2019.

38. Woods SW, Walsh BC, Powers AR, et al.: Reliability, Validity, Epidemiology, and Cultural Variation of the Structured Interview for Psychosis-Risk Syndromes (SIPS) and the Scale of Psychosis-Risk Symptoms (SOPS); in Handbook of Attenuated Psychosis Syndrome Across Cultures. Edited by Li H, Shapiro DI, Seidman LJ. Cham, Springer International Publishing, 2019 [cited 2026 Feb 17]. Available from: https://link.springer.com/10.1007/978-3-030-17336-4_5

39. Addington J, Liu L, Brummitt K, et al.: North American Prodrome Longitudinal Study (NAPLS 3): Methods and baseline description. Schizophrenia Research 243: 262–267, 2022.

40. Kroenke K, Spitzer RL, Williams JBW: The PHQ-9: Validity of a brief depression severity measure. J Gen Intern Med 16: 606–613, 2001.

41. Schafer JL, Yucel RM: Computational strategies for multivariate linear mixed-effects models with missing values. Journal of Computational and Graphical Statistics 11: 437–457, 2002.

42. Olfson M, Mojtabai R, Sampson NA: Dropout from outpatient mental health care in the United States. Psychiatric Services 60: 898–907, 2009.

43. Parker SK, Mulligan LD, Milner P, et al.: Metacognitive Therapy for Individuals at High Risk of Developing Psychosis: A Pilot Study. Front Psychol 10: 2741, 2020.

44. Mei C: Preventive interventions for individuals at ultra high risk for psychosis: An updated and extended meta-analysis. Clin Psychol Rev 86: 102005, 2021.

45. Seitz-Holland J, Jacobs GR, Reinen J, et al.: Incidence, Prevalence, and Stability of Remission in Individuals With Clinical High Risk for Psychosis. JAMA Netw Open 8: e2525644, 2025.

46. Worthington MA, Miklowitz DJ, O’Brien M, et al.: Selection for psychosocial treatment for youth at clinical high risk for psychosis based on the North American Prodrome Longitudinal Study individualized risk calculator. Early Intervention Psych 15: 96–103, 2021.

47. West ML, Parrish EM, FriedmanLYakoobian M: Treatment outcomes for young people at clinical high risk for psychosis: Data from a specialized clinic. Early Intervention Psych 16: 500–508, 2022.

48. Dohrenwend BS, Askenasy AR, Krasnoff L, et al.: Exemplification of a Method for Scaling Life Events: The PERI Life Events Scale. Journal of Health and Social Behavior 19: 205, 1978.

49. Janssen I, Krabbendam L, Bak M, et al.: Childhood abuse as a risk factor for psychotic experiences. Acta Psychiatr Scand 109: 38–45, 2004.

50. Cornblatt BA, Auther AM, Niendam T, et al.: Preliminary Findings for Two New Measures of Social and Role Functioning in the Prodromal Phase of Schizophrenia. Schizophrenia Bulletin 33: 688–702, 2007.

