## Supplementary material for "Stepped care for young people at clinical high risk for psychosis: a real-world study": Online Supplement

**Stepped care for youths at clinical high risk for psychosis: a real-world study – Online supplement**

*Implementation Details*

The PRIME Clinic treatment program was staffed by three licensed masters' level psychotherapists supervised by PRIME's clinical director, a clinical psychologist and field expert in CHR assessment, certified SIPS trainer, and contributor to the development of the specific stepped care model employed at PRIME. Each therapist received training in the SIPS, Brief Supportive Psychoeducation, P-CBT-EP, and each of the individual NAPLS Risk Calculator assessments before beginning to work with participants. Therapist inter-rater reliability on the SIPS was determined through the SIPS certification process and cases were discussed in both individual and group supervision, alongside weekly clinical team meetings. Therapists also received didactic education on the psychosis risk state and both weekly group and individual supervision with the clinical director throughout implementation. Stepping decisions, treatment approaches, and intervention fidelity were discussed in supervision with the clinical director and in weekly clinical rounds with the medical director, a licensed psychiatrist with expertise in treating young people at CHR. Participants were not billed for services, as staffing and clinical space was funded through the SAMHSA grant as well as supplementary funding from the Connecticut Department of Mental Health and Addiction Services.

Medication decision making and evaluation were performed by the PRIME Clinic's medical director and a team of psychiatry residents and fellows. Factors which influenced medication guidance, including prescription of antipsychotics outside of Step Three, were whether 1) APS severely impacted functioning or increased risk for harm to self or others, such as with command hallucinations involving consideration of self-harm or persistent paranoia leading to sustained school avoidance and 2) rapid trajectory toward conversion to psychosis, including recent increases in conviction or action based on delusional beliefs or perceptual abnormalities. Remission and adverse effects were monitored through monthly appointments with psychiatrists and regularly scheduled evaluations.

No other treatment was offered at PRIME outside of this stepped care model. In the state of Connecticut, alternative care options in the community, not specified to psychosis-spectrum concerns, included community mental health clinics and private practice options.

Treatment was adapted to address structural barriers experienced by our highly diverse patient population. Virtual sessions were offered such that patients were not limited by access to transportation and family availability. Some clinicians offered hours outside of the standard workday to further accommodate these needs. Many adolescent participants were able to access the remote sessions as a part of their school day through accommodations established by collaboration between the PRIME Clinic and local school providers. Additionally, our clinic did not require insurance and had no fee structure due to federal grant funding. As such, participants with lower economic resources, including under- or uninsured participants, were able to receive our care.

At discharge, participants would indicate to their clinical team whether they wanted continued psychopharmacological or psychotherapeutic care, and participants were not formally discharged until they had a corresponding referral with a scheduled intake. Post-PRIME referrals were tailored to the client’s personal preferences for care and relevant comorbidities (examples included speciality eating disorder or obsessive-compulsive disorder care).

Given that our therapists received substantial training to deliver this treatment model, it may be inaccessible to lower-resourced community based mental health programs. Thus, future research should prioritize the development and evaluation of virtual training materials on psychosis risk, evidence-based treatments, and risk calculator assessments. Further, a hub-and-spoke assessment and consultation model may help to implement specialized care into lower-resourced and non-specialty clinical settings, though research is needed to determine the effectiveness of this approach in psychosis risk intervention.

*NAPLS Risk Calculator Protocol*

The NAPLS Risk Calculator was administered to each participant at each timepoint by their assigned psychotherapist. The calculator utilizes six predictors: age, two of the five attenuated positive symptoms measured by the Structured Interview for Psychosis Risk Syndromes (4) (SIPS; unusual thought content [P1] and suspiciousness [P2]), neurocognitive functioning (Symbol Coding from the Brief Assessment of Cognition in Schizophrenia (39), Hopkins Verbal Learning Test Revised (40)), undesirable life events checklist (48), childhood trauma questionnaire (49), social functioning (50), and family history of psychosis. All measures are required to produce a risk calculator score. This calculator is web-based, freely available to the public, (<https://riskcalc.org/napls/>) and can be completed without advanced statistical training.

Table S.1. Tukey’s Pairwise Contrasts of Clinical Scores per time point

| **Outcome** | **Contrast** | **Δ** | **95% CI (Δ)** | **t(df)** | **p (Tukey)** |
| --- | --- | --- | --- | --- | --- |
| P | baseline vs 2mo | 1.26992587 | [-0.26, 2.80] | 2.39(203.64) | 0.166 |
| **P** | **baseline vs 6mo** | **3.72422061** | **[2.08, 5.37]** | **6.51(206.25)** | **<.001** |
| **P** | **baseline vs 12mo** | **7.25105965** | **[5.56, 8.94]** | **12.33(207.83)** | **<.001** |
| **P** | **baseline vs 18mo** | **8.37118348** | **[6.61, 10.13]** | **13.69(208.62)** | **<.001** |
| **P** | **baseline vs 24mo** | **10.58930854** | **[8.60, 12.58]** | **15.32(208.54)** | **<.001** |
| **P** | **2mo vs 6mo** | **2.45429474** | **[0.74, 4.17]** | **4.12(198.96)** | **<.001** |
| **P** | **2mo vs 12mo** | **5.98113379** | **[4.20, 7.76]** | **9.66(205.08)** | **<.001** |
| **P** | **2mo vs 18mo** | **7.10125761** | **[5.25, 8.96]** | **11.02(207.79)** | **<.001** |
| **P** | **2mo vs 24mo** | **9.31938267** | **[7.25, 11.39]** | **12.97(206.63)** | **<.001** |
| **P** | **6mo vs 12mo** | **3.52683904** | **[1.66, 5.39]** | **5.44(204.33)** | **<.001** |
| **P** | **6mo vs 18mo** | **4.64696287** | **[2.71, 6.59]** | **6.89(207.47)** | **<.001** |
| **P** | **6mo vs 24mo** | **6.86508793** | **[4.72, 9.01]** | **9.21(206.67)** | **<.001** |
| P | 12mo vs 18mo | 1.12012383 | [-0.80, 3.04] | 1.68(198.55) | 0.545 |
| **P** | **12mo vs 24mo** | **3.33824888** | **[1.22, 5.46]** | **4.53(199.82)** | **<.001** |
| P | 18mo vs 24mo | 2.21812506 | [0.07, 4.37] | 2.96(198.66) | 0.039 |
| G | baseline vs 2mo | 1.12142457 | [-0.47, 2.72] | 2.03(177.66) | 0.331 |
| **G** | **baseline vs 6mo** | **3.30459924** | **[1.55, 5.05]** | **5.44(179.86)** | **<.001** |
| **G** | **baseline vs 12mo** | **4.55094728** | **[2.76, 6.34]** | **7.31(180.29)** | **<.001** |
| **G** | **baseline vs 18mo** | **5.51832325** | **[3.70, 7.34]** | **8.75(180.03)** | **<.001** |
| **G** | **baseline vs 24mo** | **6.67410922** | **[4.64, 8.71]** | **9.43(181.19)** | **<.001** |
| G | 2mo vs 6mo | 2.18317467 | [0.32, 4.05] | 3.37(171.26) | 0.012 |
| **G** | **2mo vs 12mo** | **3.42952271** | **[1.47, 5.39]** | **5.04(179.83)** | **<.001** |
| **G** | **2mo vs 18mo** | **4.39689868** | **[2.43, 6.36]** | **6.45(177.46)** | **<.001** |
| **G** | **2mo vs 24mo** | **5.55268465** | **[3.38, 7.73]** | **7.35(179.44)** | **<.001** |
| G | 6mo vs 12mo | 1.24634805 | [-0.82, 3.31] | 1.74(179.69) | 0.510 |
| G | 6mo vs 18mo | 2.21372401 | [0.11, 4.32] | 3.03(181.35) | 0.033 |
| **G** | **6mo vs 24mo** | **3.36950998** | **[1.07, 5.67]** | **4.23(181.32)** | **<.001** |
| G | 12mo vs 18mo | 0.96737597 | [-1.12, 3.05] | 1.34(175.18) | 0.763 |
| G | 12mo vs 24mo | 2.12316194 | [-0.12, 4.37] | 2.73(172.10) | 0.075 |
| G | 18mo vs 24mo | 1.15578597 | [-1.10, 3.42] | 1.47(172.55) | 0.681 |
| N | baseline vs 2mo | 0.92201173 | [-0.96, 2.81] | 1.41(171.41) | 0.720 |
| **N** | **baseline vs 6mo** | **3.06194035** | **[0.99, 5.13]** | **4.26(172.91)** | **<.001** |
| **N** | **baseline vs 12mo** | **4.85621765** | **[2.73, 6.98]** | **6.59(173.19)** | **<.001** |
| **N** | **baseline vs 18mo** | **5.73952183** | **[3.59, 7.89]** | **7.70(172.85)** | **<.001** |
| **N** | **baseline vs 24mo** | **7.82579150** | **[5.41, 10.24]** | **9.35(173.48)** | **<.001** |
| N | 2mo vs 6mo | 2.13992863 | [-0.06, 4.34] | 2.81(165.02) | 0.062 |
| **N** | **2mo vs 12mo** | **3.93420593** | **[1.62, 6.25]** | **4.90(172.04)** | **<.001** |
| **N** | **2mo vs 18mo** | **4.81751010** | **[2.50, 7.14]** | **5.98(170.07)** | **<.001** |
| **N** | **2mo vs 24mo** | **6.90377977** | **[4.33, 9.48]** | **7.74(171.48)** | **<.001** |
| N | 6mo vs 12mo | 1.79427730 | [-0.65, 4.24] | 2.11(171.85) | 0.285 |
| N | 6mo vs 18mo | 2.67758148 | [0.19, 5.17] | 3.10(173.07) | 0.027 |
| **N** | **6mo vs 24mo** | **4.76385114** | **[2.05, 7.48]** | **5.05(172.91)** | **<.001** |
| N | 12mo vs 18mo | 0.88330418 | [-1.57, 3.34] | 1.04(168.22) | 0.905 |
| N | 12mo vs 24mo | 2.96957384 | [0.33, 5.61] | 3.24(165.63) | 0.018 |
| N | 18mo vs 24mo | 2.08626967 | [-0.58, 4.75] | 2.26(166.03) | 0.217 |
| D | baseline vs 2mo | -0.41288350 | [-1.50, 0.67] | -1.09(174.11) | 0.883 |
| D | baseline vs 6mo | 0.37966862 | [-0.81, 1.57] | 0.92(176.63) | 0.942 |
| **D** | **baseline vs 12mo** | **1.44576792** | **[0.22, 2.67]** | **3.40(177.12)** | **0.010** |
| D | baseline vs 18mo | 1.40166328 | [0.16, 2.64] | 3.26(176.87) | 0.017 |
| D | baseline vs 24mo | 2.03671985 | [0.65, 3.43] | 4.22(178.23) | <.001 |
| D | 2mo vs 6mo | 0.79255211 | [-0.48, 2.07] | 1.79(167.34) | 0.473 |
| **D** | **2mo vs 12mo** | **1.85865141** | **[0.52, 3.19]** | **4.01(176.79)** | **0.001** |
| **D** | **2mo vs 18mo** | **1.81454678** | **[0.47, 3.15]** | **3.90(174.17)** | **0.002** |
| **D** | **2mo vs 24mo** | **2.44960335** | **[0.97, 3.93]** | **4.75(176.42)** | **<.001** |
| D | 6mo vs 12mo | 1.06609930 | [-0.34, 2.48] | 2.18(176.65) | 0.254 |
| D | 6mo vs 18mo | 1.02199466 | [-0.41, 2.46] | 2.05(178.52) | 0.318 |
| D | 6mo vs 24mo | 1.65705123 | [0.09, 3.22] | 3.05(178.53) | 0.031 |
| D | 12mo vs 18mo | -0.04410463 | [-1.47, 1.38] | -0.09(171.65) | 1.000 |
| D | 12mo vs 24mo | 0.59095193 | [-0.94, 2.12] | 1.11(168.27) | 0.876 |
| D | 18mo vs 24mo | 0.63505657 | [-0.91, 2.18] | 1.19(168.76) | 0.843 |
| **PHQ-9** | **baseline vs 2mo** | **3.74633006** | **[1.61, 5.88]** | **5.04(196.78)** | **<.001** |
| **PHQ-9** | **baseline vs 6mo** | **4.65346071** | **[2.42, 6.89]** | **6.00(199.02)** | **<.001** |
| **PHQ-9** | **baseline vs 12mo** | **7.29350323** | **[4.96, 9.62]** | **9.01(201.53)** | **<.001** |
| **PHQ-9** | **baseline vs 18mo** | **7.45964335** | **[5.01, 9.90]** | **8.78(202.28)** | **<.001** |
| **PHQ-9** | **baseline vs 24mo** | **8.52657466** | **[5.76, 11.29]** | **8.88(202.14)** | **<.001** |
| PHQ-9 | 2mo vs 6mo | 0.90713065 | [-1.37, 3.18] | 1.15(189.57) | 0.860 |
| **PHQ-9** | **2mo vs 12mo** | **3.54717317** | **[1.14, 5.96]** | **4.23(197.97)** | **<.001** |
| **PHQ-9** | **2mo vs 18mo** | **3.71331329** | **[1.20, 6.23]** | **4.25(198.28)** | **<.001** |
| **PHQ-9** | **2mo vs 24mo** | **4.78024460** | **[1.96, 7.60]** | **4.88(198.50)** | **<.001** |
| PHQ-9 | 6mo vs 12mo | 2.64004252 | [0.17, 5.11] | 3.08(195.70) | 0.028 |
| PHQ-9 | 6mo vs 18mo | 2.80618264 | [0.24, 5.37] | 3.15(195.75) | 0.023 |
| **PHQ-9** | **6mo vs 24mo** | **3.87311395** | **[1.00, 6.75]** | **3.87(197.34)** | **0.002** |
| PHQ-9 | 12mo vs 18mo | 0.16614012 | [-2.41, 2.75] | 0.19(189.59) | 1.000 |
| PHQ-9 | 12mo vs 24mo | 1.23307142 | [-1.64, 4.10] | 1.24(191.06) | 0.818 |
| PHQ-9 | 18mo vs 24mo | 1.06693131 | [-1.88, 4.01] | 1.04(191.61) | 0.903 |

Bolded contrasts are significant at p<0.01

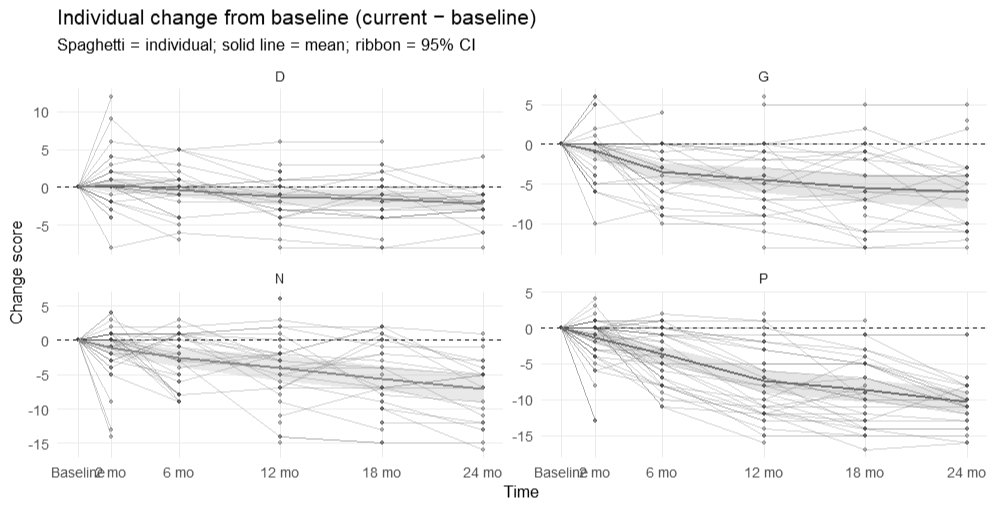

**Figure S.1. Individual level change scores.** Individual lines represent single participant symptom trajectories to final visit, emboldened lines reflect mean symptom trajectories for positive (P), negative (N), general (G), and disorganized (D) symptoms.

*Alt text:* Four-panel spaghetti plot showing individual and mean change score trajectories declining over 24 months across APS domains.

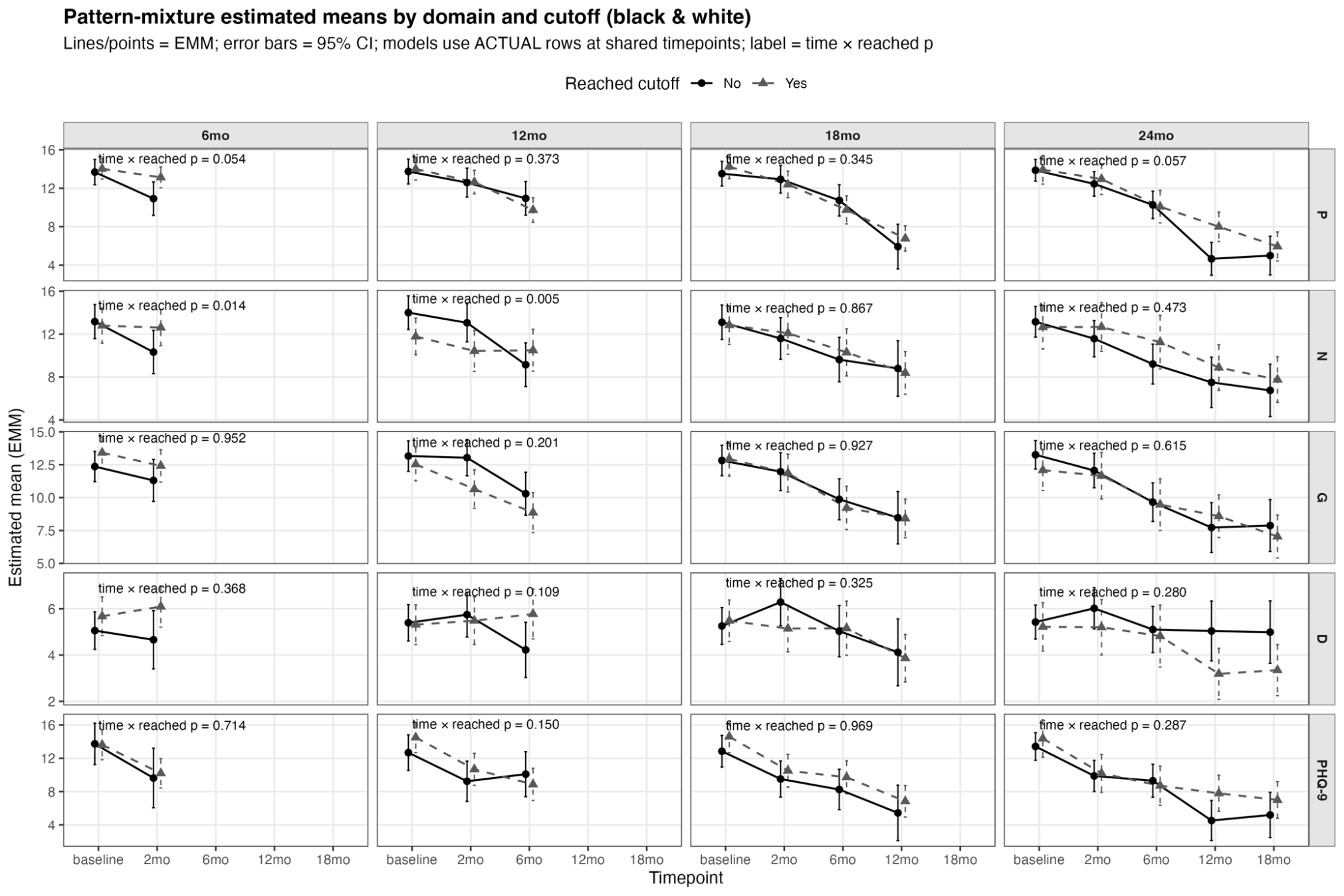

**Figure S.2. Pattern mixture analyses per point of disengagement.**  Lines and points are estimated marginal means. Ribbons are 95% confidence intervals. Panels use shared timepoints for the interaction tests of positive (P), negative (N), general (G), disorganized (D), and depressive (PHQ-9) symptoms. Lines marked with a triangle indicate individuals who completed assessment at a given time point, lines marked by circles indicate individuals who did not complete a given assessment

*Alt text:* Grid of 20 panels comparing symptom trajectories by disengagement timepoint, with interaction p-values showing mostly nonsignificant difference
